# Discovery of a Novel Missense Mutation in the RIMS1 Gene Potentially Enhances the Severity of Retinitis Pigmentosa (RP) Caused by RP1 Mutation in Humans

**DOI:** 10.1101/2024.11.01.24316231

**Authors:** José M. Lazaro-Guevara, Karen M. Garrido-Lopez, Laura Sofía Reyna Soberanis, Maria A. Sandoval-Vargas, Bryan-Josué Flores-Robles, José Luis Téllez-Arreola

## Abstract

Retinitis pigmentosa (RP) is a genetically diverse disorder characterized by the progressive degeneration of photoreceptors, ultimately leading to vision impairment and potential blindness. Understanding the disease progression and developing effective therapies is challenging due to its complex genetic landscape. This study unveils a di-genic complexity in RP involving a novel missense mutation in the RIMS1 and RP1 genes, traditionally associated with Cone-Rod Dystrophy. This mutation potentially enhances the RP phenotype, particularly in cases caused by RP1 mutations. We conducted a comprehensive genetic analysis on a family with a severe form of RP, focusing on the combined effects of RIMS1 and RP1. Using a targeted gene panel of 322 inherited retinal dystrophy (IRD) genes, we discovered a significant interaction between the RIMS1 variant and RP1 mutation within the cohort. Interestingly, patients with identical mutations exhibited substantial disease severity and progression differences. This discrepancy was particularly apparent in Patient E_1, who experienced rapid vision decline, emphasizing the impact of the mutation when combined with RP1. Biological network analysis shed light on the intricate genetic interplay, indicating a complex mechanism of disease modulation. Our findings contribute to a more nuanced understanding of RP’s genetic heterogeneity. The RIMS1 variant may serve as a modifier of the disease phenotype. This discovery expands our comprehension of the genetic factors influencing RP and underscores the importance of considering digenic interactions in future research and therapy development for retinal dystrophies.

## Introduction

Retinitis pigmentosa (RP) is a progressive degeneration of photoreceptors that leads to visual impairment and blindness. The genetic landscape of RP is diverse, involving numerous genes that contribute to the disease’s complexity and varied clinical manifestations (Lázaro-Guevara et al., 2023). RIMS1 is associated with Cone-Rod Dystrophy 2 and Cone-Rod Dystrophy 7, conditions that primarily affect cone and rod photoreceptors and lead to blindness (Miki et al., 2007; Martin-Gutierrez et al., 2022). However, emerging evidence suggests a broader role for RIMS1 in retinal diseases, including its potential involvement in different forms of RP (Weston et al., 2022; Weston et al., 2023). The discovery of a novel missense mutation in RIMS1, c.904G>C p.Glu302Gln, provides insight into the complex genetic interplay underlying retinal dystrophies. This mutation, located within a genomic region implicated in both autosomal dominant Cone-Rod Dystrophy (CORD7) and autosomal recessive RP (RP25), raises the possibility that RIMS1 may act as a phenotype enhancer in the context of other retinal dystrophy genes, particularly RP1 (Barragan et al., 2005; Lázaro-Guevara et al., 2018). To explore this hypothesis, we conducted a comprehensive genetic analysis focusing on the interaction between RIMS1 and RP1 within a pedigree exhibiting an aggressive form of RP. By examining the genetic makeup of affected individuals through a panel of 322 inherited retinal dystrophy (IRD) genes from RETNET, we aimed to uncover potential synergistic effects between these two genes that might explain the enhanced disease phenotype observed in one individual of this pedigree (Areblom et al., 2023). The role of genetic interactions in retinal dystrophies is not well understood, yet it is increasingly recognized as a critical factor in disease expression and progression. Previous studies have hinted at the complex interplay between different retinal dystrophy genes, suggesting that certain combinations of mutations may exacerbate disease severity (Lázaro-Guevara et al., 2018). This work seeks to elucidate the potential of RIMS1 to act as a phenotype enhancer in RP, particularly when co-occurring with mutations in RP1, thereby contributing to a more severe clinical presentation. Our investigation into the conservation of the RIMS1 missense mutation and its potential synergistic effect with RP1 mutations offers novel insights into the genetic architecture of retinal dystrophies. Through this analysis, we aim to expand our understanding of the genetic factors that influence disease phenotype and severity, potentially paving the way for targeted genetic therapies considering the multifaceted nature of gene interactions in retinal dystrophies.

## Methods

### Ethics statement

Approval was obtained from the Bioethics Committee of the Institute of Neurobiology (reference: #74), in accordance with the rules and regulations of the Society for Neuroscience: Policies on the Use of Animals and Humans in Neuroscience Research. All procedures were conducted in accordance with the tenets of the Declaration of Helsinki. Written informed consent was provided by all subjects.

### Subjects

This study focused on three individuals previously diagnosed with retinitis pigmentosa (RP), who was later identified through next-generation sequencing (NGS) as having autosomal recessive RP1 (arRP1) linked to the Ser542* mutation. The cohort consisted of two female patients and one male patient, all full siblings, as confirmed by kinship coefficient calculations. Each subject was clinically diagnosed with retinitis pigmentosa (RP) after the presence of nyctalopia (night blindness) between their twenties and thirties. This early symptom onset was followed by a progressive narrowing of their visual field, leading to concentric tunnel vision over the next two decades. Notably, the disease’s progression varied markedly among the individuals. Patient E_1 encountered total vision loss within five years following their initial symptom onset. Patients C_2 and O_3 had more evident symptoms in their late thirties and retained some degree of vision for an extended period. C_2 preserved residual vision for 15 additional years and O_3 for 20 more years, respectively. The formal diagnosis of retinitis pigmentosa in each case was made by fundus examinations, which revealed the characteristic retinal pigment deposits indicative of the condition.

### Panel Construction and Digenic Interrogation

#### Targeted Gene Panel Selection

Dissect the genetic underpinnings of aggravated retinitis pigmentosa phenotype within the cohort. To achieve this, we curated a targeted gene panel comprised of 286 genes associated with inherited retinal dystrophies (IRDs), as delineated by the exhaustive gene list in Areblom et al. (2023). We excluded mitochondrial DNA genes from our analysis due to their absence in Whole Exome Sequencing (WES) data.

#### Sequencing and Data Extraction

Our genetic analysis utilized Low-Coverage Whole Genome Sequencing (LC-WGS) and WES. To build upon our previous experience, we analyzed samples from three individuals identified in our prior 2023 research (Lázaro-Guevara et al.) affected by RP1 (Ser542*), applying refined techniques for extracting the specified gene subset from LC-WGS sequencing datasets.

#### Gene Panel Extraction

We revisited the gene panel for extraction from the same three RP1-affected patients (two females and one male), directly retrieving gene region data from aligned Binary Alignment Map (BAM) files. Variant calling was executed using the DRAGEN v.4.2 pipeline, ensuring high precision in our genetic analysis.

#### Panel Interrogation and Variant Filtering

To scrutinize the gene panel further, we employed Gene. iobio v. 4.9 (Di Sera et al., 2021) enabled us to exclude known benign variants and areas lacking adequate coverage (Fig 2). We then processed the resulting Variant Call Format (VCF) file through DiVas V1.1, applying specific Human Phenotype Ontology (HPO) terms—such as Nyctalopia (HP:0000662) and Visual impairment (HP:0000505)—to refine interactions as shown in the protocol by Taha et al. (2022). Only variants meeting a Pathogenicity Score (PS) of 0.90 or above were considered for further analysis.

**Figure 1:**
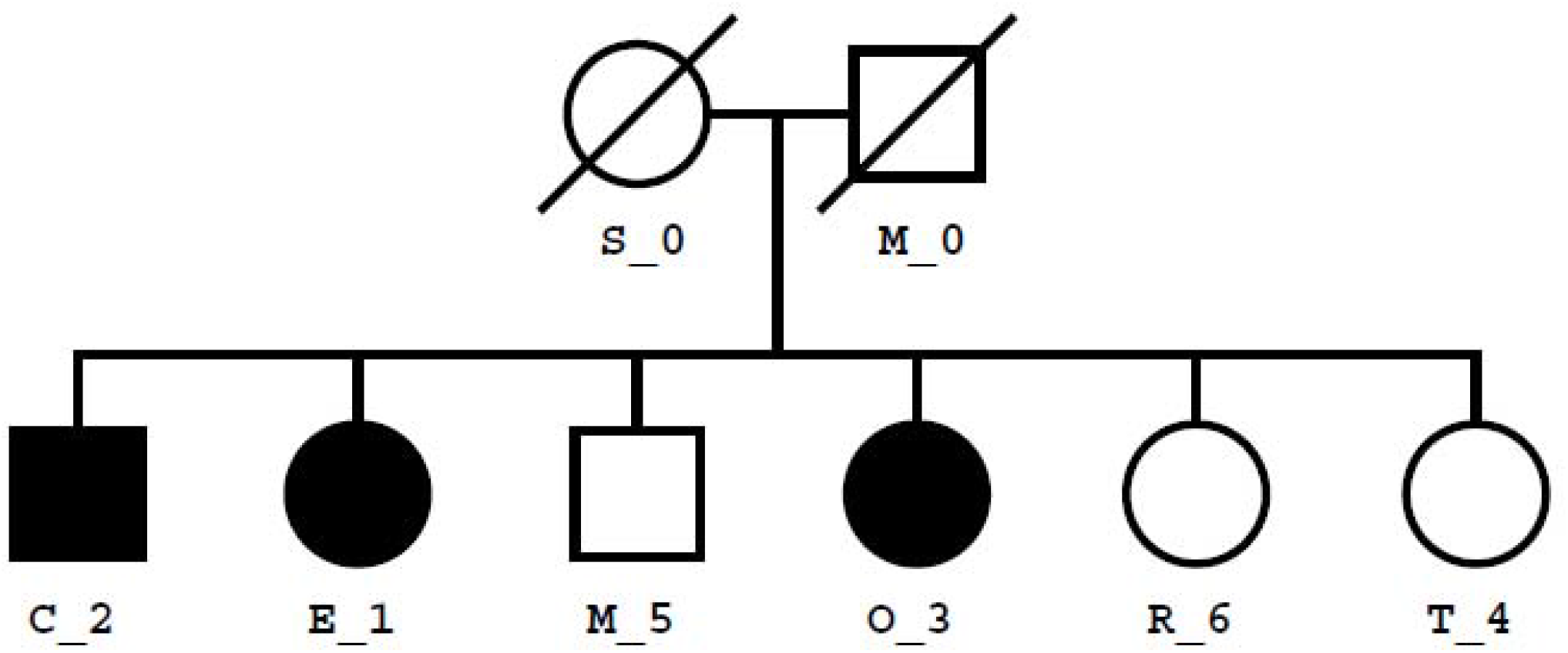
Pedigree Illustrating RP Inheritance. This pedigree outlines the inheritance pattern for the family’s retinitis pigmentosa (RP), correlating with patient identifiers used in the study. The squares represent male family members, while the circles represent female members. Filled symbols indicate individuals diagnosed with RP, and the line through a symbol denotes a deceased individual. The index patient (proband) is marked with an arrow. Notably, patient E_1 exhibits the most severe phenotype, aligning with unique genetic findings in the study. Genetic homogeneity was confirmed through principal component analysis (PCA), with all sequenced family members identified as Admixed American, and no significant consanguinity or inbreeding detected, as evidenced by kinship coefficients and genomic inbreeding coefficients (fROH).

**Figure 2:**
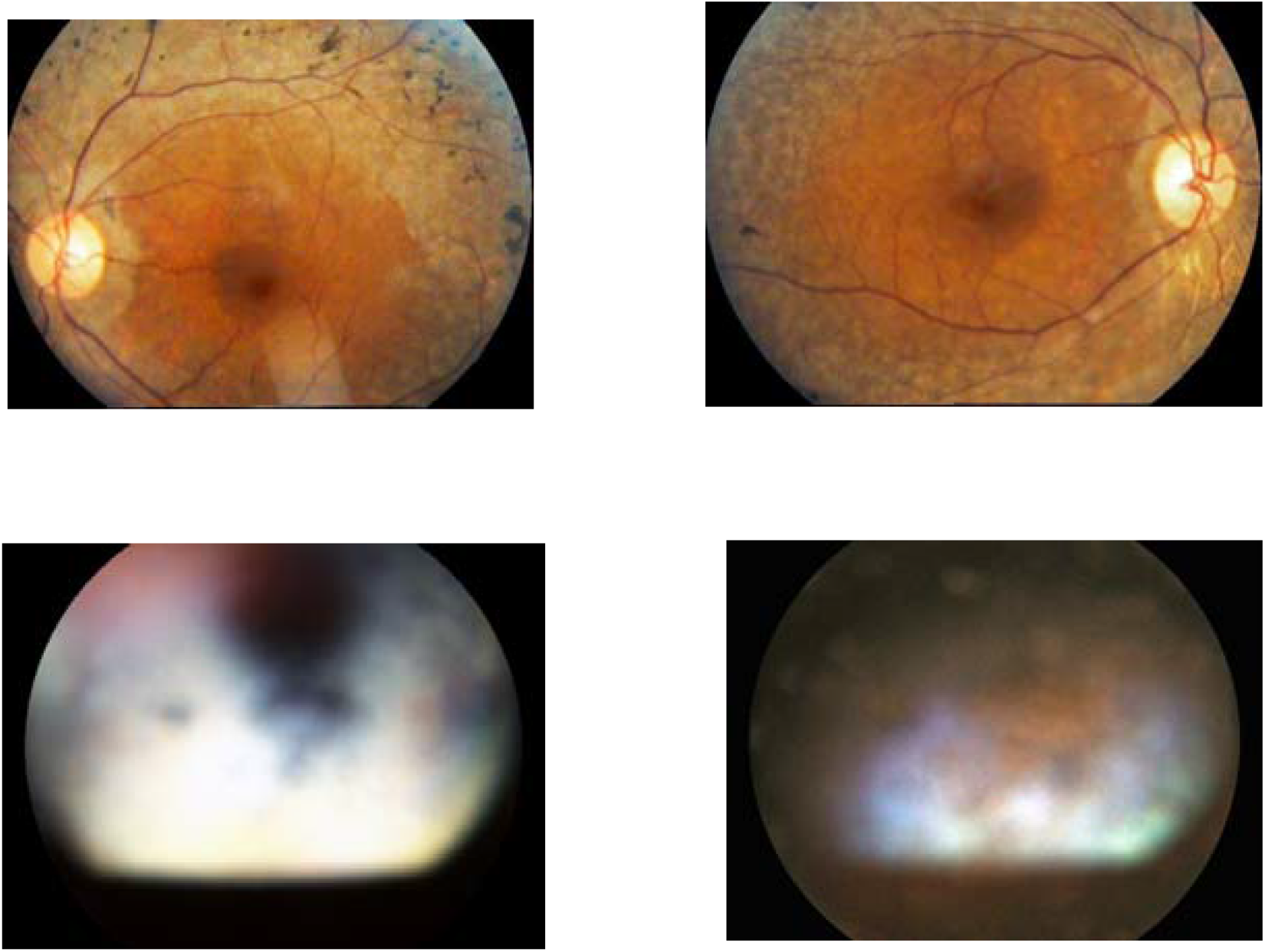
Comparative Ocular Fundus Imagery in RP1. This figure presents fundus photographs of two patients diagnosed with RP1 (both eyes), offering a visual contrast of disease manifestation. The lower panel showcases the fundus of Patient E_1, highlighting the characteristic subcapsular cataracts indicative of advanced RP1. This common complication can be identified as a cloudy, greyish discoloration at the back of the lens, obscuring the underlying retinal details. The upper panel exhibits the fundus of Patient O_3, which displays the classical signs of RP1, including spicule-like pigment deposits that resemble bone spicules, indicative of the disease’s progressive nature. These speculations are scattered throughout the retina’s periphery, reflecting the pigmentary changes associated with photoreceptor degeneration. Both images underscore the varied clinical presentation of RP1 and help delineate the disease’s progression markers.

#### Identification of Digenic Pairs

A presence/absence table constructed to identify variants co-occurring with the RP1 mutation C1625 C>G (p.Ser542*), focusing on those with the highest PS. Digenic pairs exclusively found in patient E_1, who displayed the most severe genotype, were prioritized for further investigation and annotation using CADD score and phyloP to evaluate deleterious risk and aminoacid conservation among other species.

#### Biological Networks

The resulting top 5 digenic co-occurrences examined via topological-biological networks like RPGeNet v2.0 and Genes Hubs (Lázaro-Guevara et al., 2018; Arenas-Galnares et al., 2019) to find biological connections that elucidated the interaction between the digenic findings (Fig. 3). Networks work expanded using node grade and topological layout via Cytoscape V 3.10.2. Closes node degree for RP1 and Cone rode dystrophy were selected using node centrality and closes neighboring.

**Figure 3:**
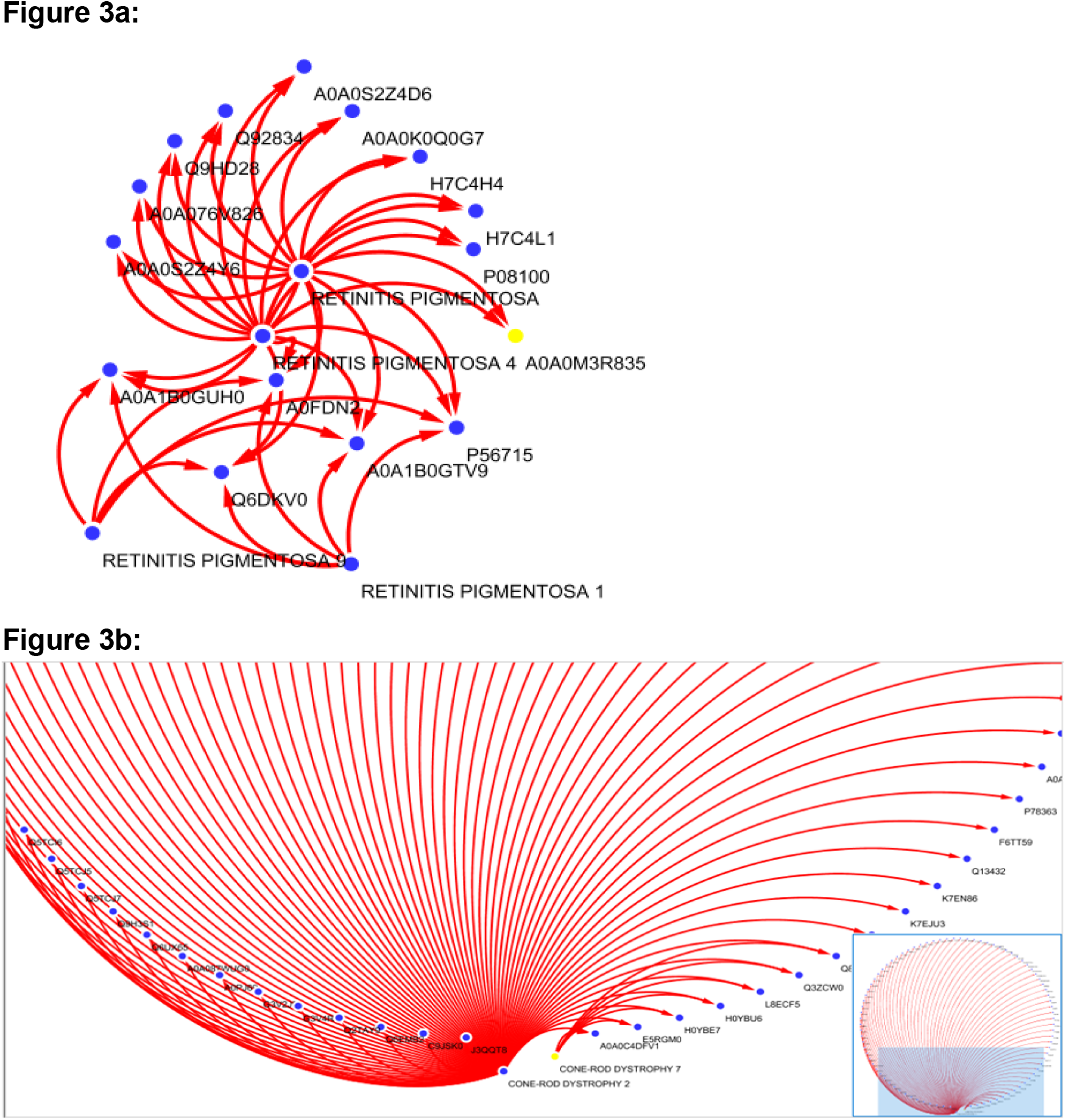
Biological Network Analysis of Retinitis Pigmentosa and Cone-Rod Dystrophy. Panels A and b demonstrate the biological networks that may underlie the digenic interactions influencing retinitis pigmentosa (RP) phenotypes, focusing on the relationships between RP1, CORD7, and other associated genetic variants. In panel a, the network captures the interconnectivity for RP types 1, 9, and 4, while panel b showcases the networks for cone-rod dystrophy types 2 and 7. The red lines denote the direct and indirect interactions within the networks, with significant nodes representing gene variants. The nodal interactions, particularly those involving RIMS1 and RP1, highlight the complex genetic interplay potentially responsible for the variability in clinical manifestations of RP. The degree of separation in these networks follows the patterns suggested by Lazaro et al. (2018), with the main interactions occurring at least two nodes apart, implying a broader genetic influence that may intensify disease phenotypes in affected individuals.

## Results

In 2023, our research introduced a three-generation pedigree, focusing on a family with clinically diagnosed retinitis pigmentosa (RP) without prior genetic confirmation. Our initial findings, which utilized both Low Coverage Whole Genome Sequencing (LC-WGS) and Whole Exome Sequencing (WES), pinpointed a rare mutation in the RP1 gene, c.1625C>G p.Ser542*, as the causative factor for the autosomal recessive RP phenotype. This discovery was corroborated by additional ClinVar reports affirming this variant’s pathogenic nature (José et al. et al., 2023). This work delves into the first generation of this RP-enriched family, depicted in Figure 1, to scrutinize the phenotypic inter variability—from milder to more aggressive forms— among family members sharing the same RP1 mutation. We uncovered a novel missense mutation in the RIMS1 gene, c.904G>C p.Glu302Gln. This mutation’s interplay with the established RP1 mutation illuminates the intricate genetic complexity of individuals afflicted with a particularly aggressive form of RP. Our comprehensive genetic analysis aimed to delineate how these and other digenic variant pairs might intensify the phenotypic severity of RP within the cohort.

### Clinical Progression of Retinitis Pigmentosa

Retinitis Pigmentosa (RP) progression among the studied patients underscores the disease’s heterogeneity. A detailed comparison of their clinical characteristics among subjects is presented in Table 1. Patient E_1 experienced a remarkably rapid decline in vision, transitioning from diagnosis at age early 20ths to complete vision loss within 5 years. This patient’s trajectory highlights a severe progression of RP, with additional complications, including macular edema and subcapsular cataracts, indicative of an aggressive disease phenotype. Patients C_2 and O_3 had gradual progression of RP, diagnosed in their early to mid-thirties, maintaining partial vision for decades, with C_2 experiencing significant vision loss by late 40s and O_3 by age middle-50ths. Notably, neither of these patients developed macular edema or subcapsular cataracts, and O_3 retained day/night perception abilities. The variability in onset, progression, and associated clinical features, as detailed in Table 1, accentuates the complexity of RP and the critical role of genetic and possibly environmental factors in influencing disease severity and progression.

**Table 1:**
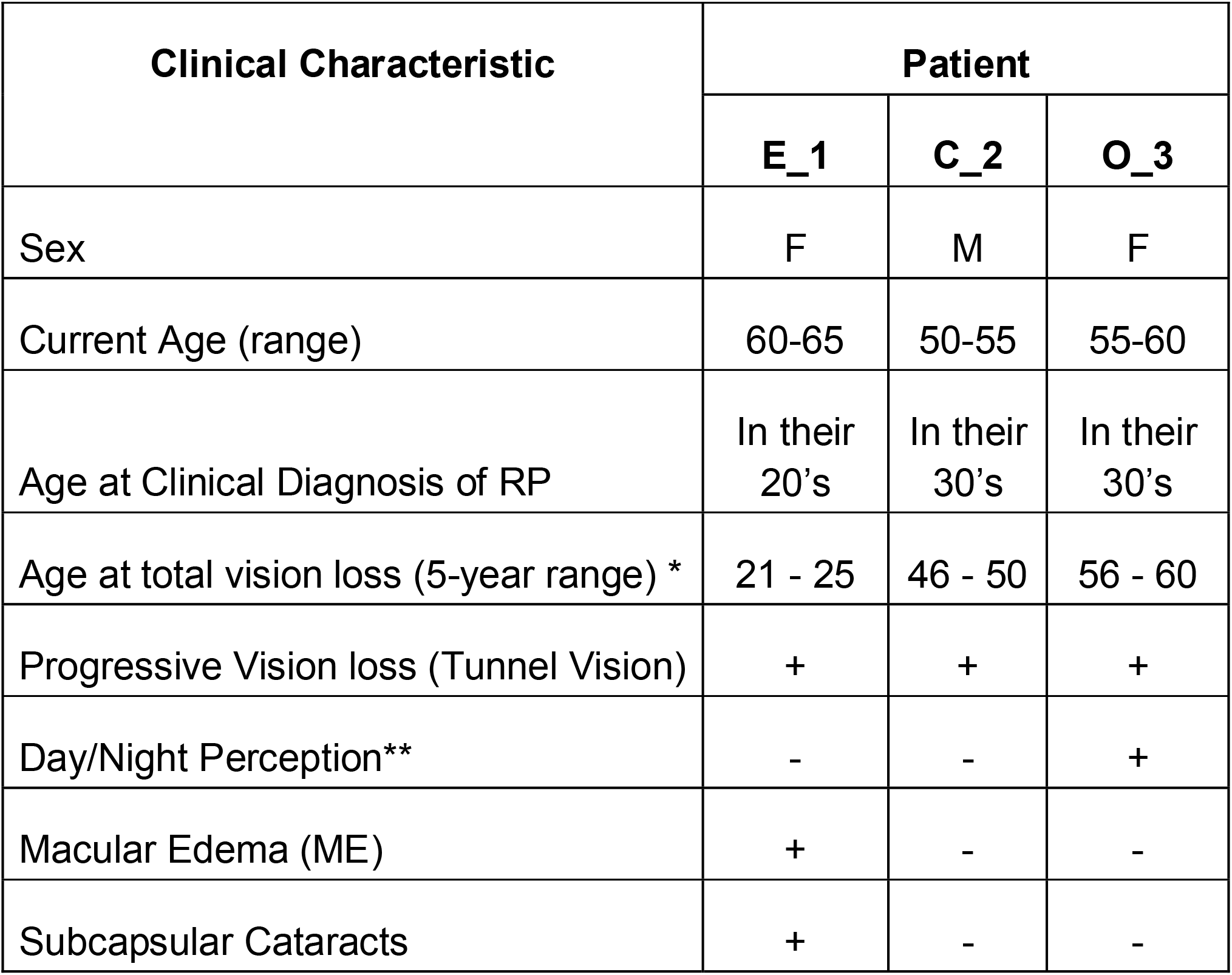
Clinical Progression of Retinitis Pigmentosa. Patient-specific progression and clinical characteristics for RP. “+” indicates presence and “-” absence of features. Patient IDs are E_1, C_2, and O_3, representing varied clinical onset and progression. *Age at total vision loss is approximate. **Day/Night perception noted only for patient O_3.

| Clinical Characteristic | Patient |  |  |
| --- | --- | --- | --- |
|  | E_1 | C_2 | O_3 |
| Sex | F | M | F |
| Current Age (range) | 60-65 | 50-55 | 55-60 |
| Age at Clinical Diagnosis of RP | In their 20's | In their 30's | In their 30's |
| Age at total vision loss (5-year range) * | 21 - 25 | 46 - 50 | 56 - 60 |
| Progressive Vision loss (Tunnel Vision) | + | + | + |
| Day/Night Perception** | - | - | + |
| Macular Edema (ME) | + | - | - |
| Subcapsular Cataracts | + | - | - |

**Table 2:** DiVas Digenic Variant Scores and Genomic Annotation. Interaction of digenic variant pairs across three RP patients, E_1, C_2, and O_3, highlighting their DiVas and CADD scores and genomic context including PhyloP, ClinVar, and gnomAD v4 frequencies. “X” marks the presence of a variant in the patient. DiVas scores close to 1 suggest strong digenic interaction potential. *PhyloP scores estimate evolutionary conservation. **ClinVar annotations provide clinical significance of variants. ***GnomAD frequencies indicate variant rarity. The RIMS1 c.904G>C variant appears exclusively with high impact in patient E_1, suggesting a novel genetic interaction in RP severity.

| No | Digenic Variant Pair | Patient |  |  | DiVas Score | CADD Score | Phylo P * | ClinVar** | gnomAD v4 *** |
| --- | --- | --- | --- | --- | --- | --- | --- | --- | --- |
|  |  | E_1 | C_2 | O_3 |  |  |  |  |  |
| 1 | TRPM1 c.1879G>A<br>p.Val627Met - RP1<br>c.1625C>G p.Ser542* | X | X | X | 1 | 23.1 - 35 | 5.92 - 8.90 | Benign - Pathogenic | 3.04 e-2 - 1.24 e-5 |
| 2 | RBP3 c.2919C>A<br>p.Ser973Ser - RP1<br>c.1625C>G p.Ser542* | X | X | X | .99999 | 0.077 - 35 | -1.84 - 8.90 | Likely Benign - Pathogenic | 3.66 e-5 - 1.24 e-5 |
| 3 | RIMS1 c.904G>C<br>p.Glu302Gln - RP1<br>c.1625C>G p.Ser542* | X | -- | -- | .99998 | 18.9 - 35 | 8.74 - 8.90 | Not Reported - Pathogenic | ----<br>1.24 e-5 |
| 4 | RIMS1 c.904G>C<br>p.Glu302Gln - TRPM1<br>c.1879G>A p.Val627Met | X | -- | -- | .99974 | 18.9 - 23.1 | 8.74 - 5.92 | Not Reported - Benign | ----<br>3.04 e-2 |
| 5 | TRPM1 c.1879G>A<br>p.Val627Met - RBP3<br>c.2919C>A p.Ser973Ser | X | X | X | .99922 | 23.1 - 0.077 | 5.92 - -1.84 | Benign - Likely Benign | 3.04 e-2 - 3.66 e-5 |

### Genetic Interactions and Variant Analysis

Our focus shifted to identifying digenic interactions that may affect the RP phenotype. The interplay between the novel RIMS1 and RP1 mutations was particularly striking, presenting exclusively in the most severely affected patient, E_1. That correlation, underscored by high DiVas and CADD scores (RIMS1 18.9 CADD score), signals a critical avenue for further investigation. The RIMS1 variant’s novelty suggests we found a genetic contributor linked to RP severity.

### Biological Network Analysis

Results of biological networks shed light on the potential mechanisms by which these digenic pairs could modulate disease phenotype. These complex interactions have been shown to occur in multiple networks (protein-protein) or (phenotype-protein) networks. The primary interaction is at least two node degrees apart (Lazaro et al., 2018), networks for Retinitis Pigmentosa 1, 9, 4, and cone-rod dystrophies 2 and 7 (figure 3). Although the interaction between RIMS1 and RP1 is unclear, mutations on both point to a sophisticated genetic landscape, potentially crucial to understanding the variability seen in clinical presentations. This digenic scenario, particularly in severely affected individuals, raises the possibility of RIMS1 acting as a phenotype enhancer, further complicating the genetic underpinnings of RP.

## Discussion

This study details a new missense mutation in the RIMS1 gene, which is proposed to enhance the symptoms of Retinitis Pigmentosa (RP) caused by mutations in the RP1 gene. Our findings contribute to understanding the genetic complexity of RP, as the variability in disease progression, even among genetically similar individuals, underscores the multifaceted nature of its development. The significant variability in RP severity, observed even among siblings sharing identical genetic mutations, highlights the disease’s heterogeneity and the limitations of predicting clinical outcomes based solely on genetic diagnosis (Berson, 2007). This variability suggests that other factors beyond single gene mutations are crucial in disease manifestation and progression. Our study focuses on the interaction of genes that cause RP, particularly RIMS1 and RP1 mutations, shedding light on the complex genetic interplay that can exacerbate disease severity. As reported in other cohort studies, the association of RP1 and EYS mutations with macular edema (ME) indicates a possible genetic predisposition for ME in RP (Arias et al., 2023). This finding adds another layer to the genetic landscape of RP, suggesting that specific genetic configurations may predispose individuals to more severe phenotypic expressions, such as ME, which can further impact visual prognosis. The enhanced retinal phenotype observed in patients carrying variants in RIMS1 and EYS aligns with previous reports of heavy pigmentation in the macular area in similar genetic configurations (Suvannaboon et al., 2022). This observation supports the hypothesis that RIMS1 variants may enhance phenotypes in RP, particularly when combined with other pathogenic mutations. This role of RIMS1, traditionally associated with Cone-Rod Dystrophies, in potentially exacerbating RP phenotypes, represents a significant expansion of our understanding of its involvement in retinal diseases. Furthermore, our exploration into the genetic complexity of Retinitis Pigmentosa (RP) is complemented by studies investigating the role of the RIMS1 gene in retinal dystrophies. Barragán et al. (2005) conducted a molecular analysis that sought to understand the involvement of RIMS1 in autosomal recessive RP (arRP), particularly in the context of RP25. Their findings suggested that while RIMS1 mutations are a known cause of autosomal dominant Cone-Rod Dystrophy (CORD7), several arRP families established a possible link between RIMS1 and the RP25 locus, That indicates the complex genetic interplay and the potential for other genetic factors or mutations to contribute to RP’s heterogeneity significantly (Barragán et al., 2005). Sisodiya et al. (2007) explored the broader implications of RIMS1 mutations on phenotypic expressions beyond their known association with Cone-Rod Dystrophies. Their investigation into a kindred with RIMS1 mutation-related retinal dystrophy and enhanced cognitive abilities highlights the diverse roles of genes implicated in retinal diseases. While their study primarily focuses on cognitive enhancement in the context of RIMS1 mutations, further research is needed into the multifaceted effects of RIMS1 mutations and their potential impact on retinal diseases, including RP (Sisodiya et al., 2007). Identification of digenic interactions through biological network analysis, such as in our study, suggests the genetic complexity landscape and points to the need for further investigation into the molecular mechanisms of RP, which could drive the development of new targets for therapeutic intervention. Our results indicate that the RIMS1 variant is pathogenic. The CADD score of 18.90, slightly below the threshold typically indicative of pathogenicity (scores above 20), nonetheless represents a significant deviation from the norm, suggesting potential deleterious effects. This score places the variant in a gray area where its impact on the protein’s function and the disease phenotype warrants closer examination. Given the critical role of RIMS1 in synaptic function and its novel association with RP in our study, the elevated CADD score further emphasizes the need for functional studies to elucidate the precise mechanism by which this variant contributes to RP severity. The findings of this study highlight the necessity for accurate and sensitive methods to track the progression of RP (Colombo et al., 2018). These methods are fundamental for future clinical trials and therapeutic strategies, particularly in a genetically and phenotypically heterogeneous disease like RP. Furthermore, the potential for targeted genetic therapies, informed by a deeper understanding of the genetic interactions contributing to RP severity, offers a promising avenue for personalized treatment approaches.

In conclusion, our study provides valuable insights into the genetic architecture of Retinitis Pigmentosa. It highlights the complex interplay between genes such as RIMS1 and RP1 and their potential to influence the disease phenotype. These findings expand our understanding of the genetic basis of RP and lay the groundwork for future research aimed at developing targeted therapies and improving disease management strategies for individuals affected by this challenging condition.

### Patient Privacy

All identifier codes (e.g. M_5) in this study are not back traceable to medical identifiers, and all the codes used as identifier were randomly generated and are unique for this study and only the researchers participating in this study had access to them.

## Supporting information

Coverage_Table

Phylo_CADD_scores_table

Divas_Run

## Data Availability

All data produced are available online at https://home.chpc.utah.edu/~u1123911/Plos&20Genetics_Data/

https://home.chpc.utah.edu/~u1123911/Plos&20Genetics_Data/Graphical&20Abstract.jpg

## Acknowledgments

This work was supported by the Department of Rheumatology from San Pedro Hospital in Logroño and the SCT CF-2019-1759 CONACYT grant. The computational calculations and graphics in this research were partly enabled by support from the Digital Research Alliance of Canada (alliancecan.ca) RRG grant. We acknowledge Dr. Stéphanie Thébault’s support in providing critical comments to the manuscript for this research.

---

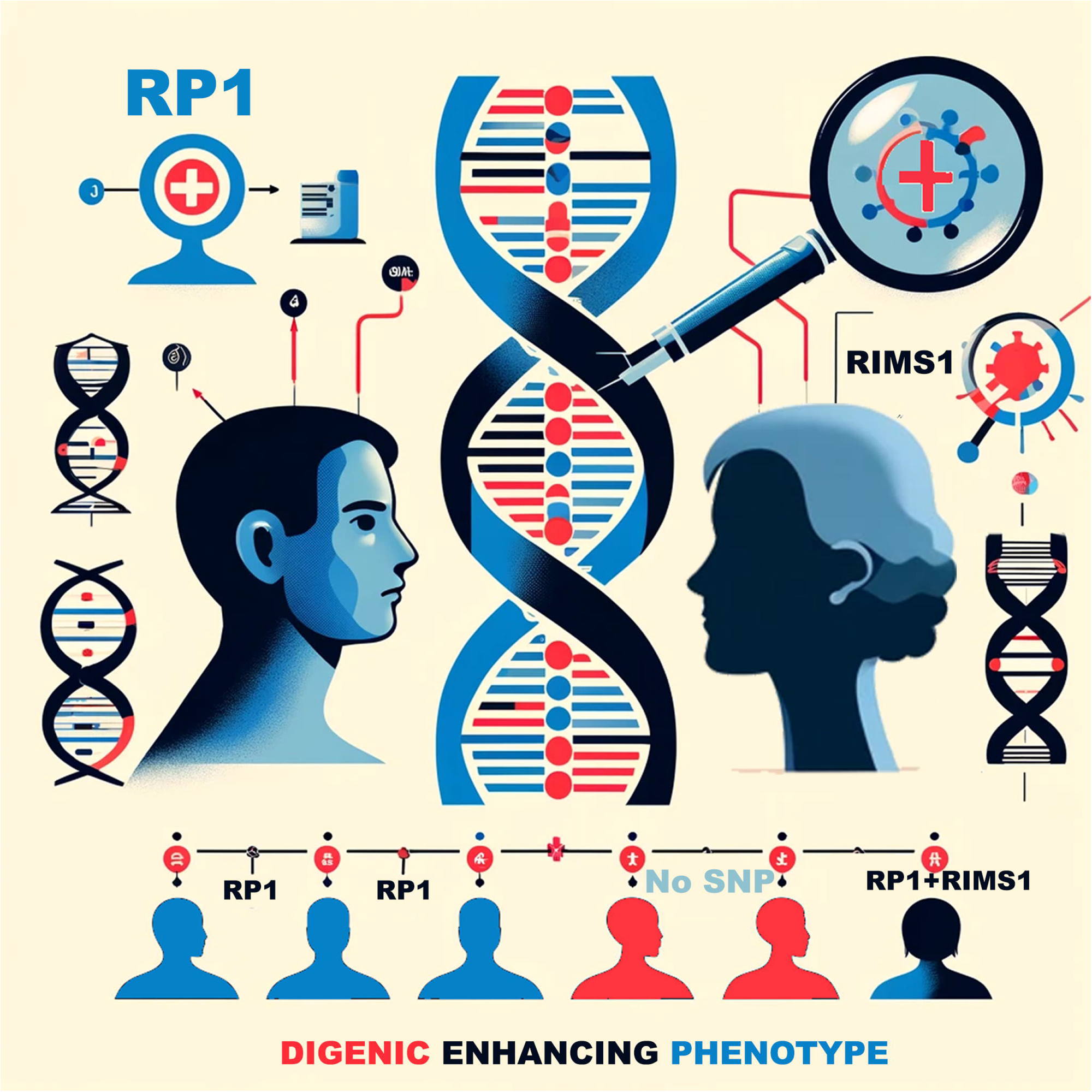

