## Supplementary figures and images for "Discovery of a Novel Missense Mutation in the RIMS1 Gene Potentially Enhances the Severity of Retinitis Pigmentosa (RP) Caused by RP1 Mutation in Humans"

### Coverage_Table

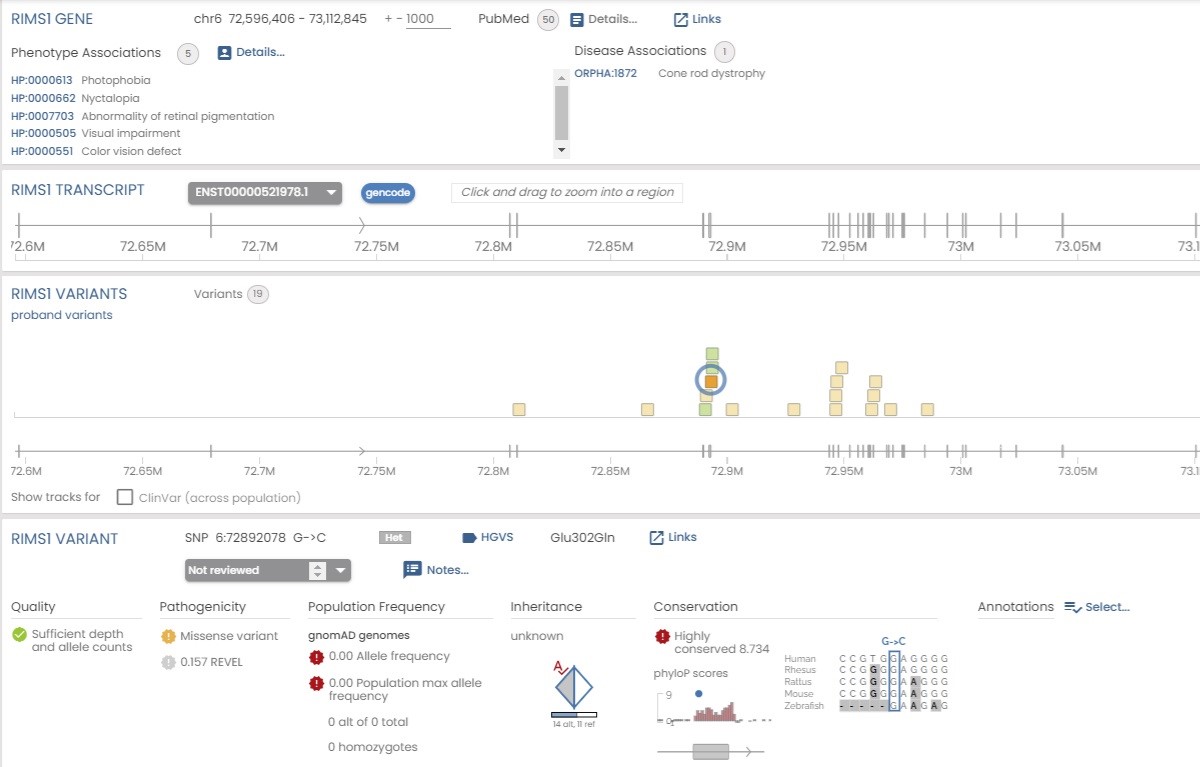
