## Supplementary material for "Discovery of a Novel Missense Mutation in the RIMS1 Gene Potentially Enhances the Severity of Retinitis Pigmentosa (RP) Caused by RP1 Mutation in Humans": Divas_Run

### DIVAs report

Powered by enGenome

#### GENERAL INFORMATION

| | |
|---|---|
| DIVAs version | DIVAs v1.1 |

#### HPO TERMS

| HPO TERM | DESCRIPTION |
|---|---|
| HP:0000662 | Nyctalopia |
| HP:0000505 | Visual impairment |
| HP:0007703 | Abnormality of retinal pigmentation |
| HP:0007947 | Pericentral retinitis pigmentosa |
| HP:0000546 | Retinal degeneration |

#### FAMILY HISTORY

| Sample | Father | Mother | Sex | Affected |
|---|---|---|---|---|
| | | | | Yes |

#### ANALYSIS PARAMETERS

| | | |
|---|---|---|
| QUALITY | 30.0 | Only variants with QUALITY >= of this threshold will be retained |
| GENOTYPE QUALITY | 0.0 | Only variants with GENOTYPE QUALITY >= of this threshold will be retained |
| FILTER | PASS | Only variants with FILTER field with this value will be retained |
| PATHOGENICITY SCORE | 1.0 | Only variants with PS >= of this threshold will be retained |

### STATISTICS

| | | |
|---|---|---|
| TOTAL VARIANTS | 229267 | Variants present in the original VCF |
| RETAINED VARIANTS | 1899 | Variants with QUALITY >= 30.0, GENOTYPE QUALITY >= 0.0, FILTER=PASS and PS >= 1.0 |
| TOTAL COMBINATIONS | 111639 | Number of digenic combinations evaluated by DIVAs |
| NUMBER OF PATHOGENIC COMBINATIONS | 2859 | Number of digenic combinations classified pathogenic by DIVAs |

### RESULTS

| Gene A | HGVS.c A | HGVS.p A | Effect A | Gene B | HGVS.c B | HGVS.p B | Effect B | Score |
|---|---|---|---|---|---|---|---|---|
| TRPM1 | c.1879G>A | p.Val627Met | missense | RP1 | c.1625C>G | p.Ser542* | stop gained | 1.00000 |
| RBP3 | c.2919C>A | p.Ser973Ser | synonymous | RP1 | c.1625C>G | p.Ser542* | stop gained | 1.00000 |
| CNGB1 | c.874+2074G>T; c.291-145A>C | .; . | intron; intron | RP1 | c.1625C>G | p.Ser542* | stop gained | 0.99999 |
| VCAN | c.*386G>A | . | 3 prime UTR | RP1 | c.1625C>G | p.Ser542* | stop gained | 0.99999 |
| BEST1 | c.*1721G>A; c.*1692G>A | .; . | 3 prime UTR; 3 prime UTR | RP1 | c.1625C>G | p.Ser542* | stop gained | 0.99999 |
| PRPH2 | c.582-4734_582-4733insGTTT | . | intron | RP1 | c.1625C>G | p.Ser542* | stop gained | 0.99999 |
| PROM1 | c.1683-1269_1683-1267delGTC; c.1683-1283_1683-1275delCCTTCTTCT | .; . | intron; intron | RP1 | c.1625C>G | p.Ser542* | stop gained | 0.99998 |
| RIMS1 | c.904G>C | p.Glu302Gln | missense | RP1 | c.1625C>G | p.Ser542* | stop gained | 0.99998 |
| VCAN | c.*386G>A | . | 3 prime UTR | TRPM1 | c.1879G>A | p.Val627Met | missense | 0.99998 |
| TULP1 | c.823-754_823-750delTACAC | . | intron | RP1 | c.1625C>G | p.Ser542* | stop gained | 0.99998 |
| CHM | c.1244+97A>T | . | intron | RP1 | c.1625C>G | p.Ser542* | stop gained | 0.99997 |

| | | | | | | | | |
|---|---|---|---|---|---|---|---|---|
| ERCC3 | c.1945+1915_1945+1918delAAAA | . | intron | RP1 | c.1625C>G | p.Ser542* | stop gained | 0.99997 |
| POC1B | c.1114-12674_1114-12672delTTT | . | intron | RP1 | c.1625C>G | p.Ser542* | stop gained | 0.99996 |
| AHR | c.2404-73T>C | . | intron | RP1 | c.1625C>G | p.Ser542* | stop gained | 0.99993 |
| GPR143 | c.250+1044_250+1063delTGTGTGTGTGTGTGTGTGTG | . | intron | RP1 | c.1625C>G | p.Ser542* | stop gained | 0.99993 |
| ZNF141 | c.204_205insGA; c.209_210delTC | p.Lys69fs; p.Ile70fs | frameshift; frameshift | RP1 | c.1625C>G | p.Ser542* | stop gained | 0.99992 |
| ERCC4 | c.1812-541G>A | . | intron | RP1 | c.1625C>G | p.Ser542* | stop gained | 0.99989 |
| VCAN | c.*386G>A | . | 3 prime UTR | BEST1 | c.*1721G>A; c.*1692G>A | .; . | 3 prime UTR; 3 prime UTR | 0.99988 |
| PRIMPOL | c.-59-7delG | . | splice region, intron | RP1 | c.1625C>G | p.Ser542* | stop gained | 0.99988 |
| POLR3A | c.3472C>T | p.Leu1158Phe | missense | RP1 | c.1625C>G | p.Ser542* | stop gained | 0.99986 |
| DHX38 | c.3477+5G>A; c.*246T>G | .; . | splice region, intron; 3 prime UTR | RP1 | c.1625C>G | p.Ser542* | stop gained | 0.99983 |
| LPL | c.344C>A; c.875G>A | p.Ser115*; p.Ser292Asn | stop gained; missense | RP1 | c.1625C>G | p.Ser542* | stop gained | 0.99980 |
| FTH1 | c.541_543delAAT; c.388-1_388insATCCCCAC | p.Asn181del; . | conservative inframe deletion; splice acceptor, intron | RP1 | c.1625C>G | p.Ser542* | stop gained | 0.99979 |

| | | | | | | | | |
|---|---|---|---|---|---|---|---|---|
| DHX37 | c.2045+1648T>C | . | intron | RP1 | c.1625C>G | p.Ser542* | stop gained | 0.99978 |
| BEST1 | c.*1721G>A; c.*1692G>A | .; . | 3 prime UTR; 3 prime UTR | RBP3 | c.2919C>A | p.Ser973Ser | synonymous | 0.99977 |
| GRIK2 | c.899G>A | p.Arg300Gln | missense | RP1 | c.1625C>G | p.Ser542* | stop gained | 0.99976 |
| SDCCAG8 | c.1744+124C>T | . | intron | RP1 | c.1625C>G | p.Ser542* | stop gained | 0.99976 |
| RIMS1 | c.904G>C | p.Glu302Gln | missense | POC1B | c.1114-12674_1114-12672delTTT | . | intron | 0.99976 |
| RIMS1 | c.904G>C | p.Glu302Gln | missense | TRPM1 | c.1879G>A | p.Val627Met | missense | 0.99974 |
| COL3A1 | c.4011+1G>T; c.3997G>A | .; p.Asp1333Asn | splice donor, intron; missense | RP1 | c.1625C>G | p.Ser542* | stop gained | 0.99963 |
| CNBP | c.*458C>A | . | 3 prime UTR | RP1 | c.1625C>G | p.Ser542* | stop gained | 0.99960 |
| HTT | c.102delG; c.99_100delGC | p.Gln34fs; p.Gln34fs | frameshift; frameshift | RP1 | c.1625C>G | p.Ser542* | stop gained | 0.99955 |
| POC1B | c.1114-12674_1114-12672delTTT | . | intron | TRPM1 | c.1879G>A | p.Val627Met | missense | 0.99955 |
| ARMC2 | c.1024-3233G>A | . | intron | RP1 | c.1625C>G | p.Ser542* | stop gained | 0.99954 |
| CPE | c.41G>C; c.1113_1113+1insATACACCGAGGTGTTAAAGGGTTTGTCCGTGACCTTCAGG | p.Gly14Ala; . | missense; splice donor, intron | RP1 | c.1625C>G | p.Ser542* | stop gained | 0.99954 |
| DNAAF4 | c.862_866delAAGAA | p.Lys288fs | frameshift | RP1 | c.1625C>G | p.Ser542* | stop gained | 0.99951 |
| TULP1 | c.823-754_823-750delTACAC | . | intron | BEST1 | c.*1721G>A; c.*1692G>A | .; . | 3 prime UTR; 3 prime UTR | 0.99950 |

| | | | | | | | | |
|---|---|---|---|---|---|---|---|---|
| RB1 | c.607+2970T>C | . | intron | RP1 | c.1625C>G | p.Ser542* | stop gained | 0.99950 |
| BBS9 | c.2116-30675dupA | . | intron | RP1 | c.1625C>G | p.Ser542* | stop gained | 0.99947 |
| CDH2 | c.*804T>A | . | 3 prime UTR | RP1 | c.1625C>G | p.Ser542* | stop gained | 0.99946 |
| DDX3X | . | . | intron | RP1 | c.1625C>G | p.Ser542* | stop gained | 0.99946 |
| TFAP2A | c.52-1352G>A | . | intron | RP1 | c.1625C>G | p.Ser542* | stop gained | 0.99945 |
| GJA1 | c.*1119_*1120insA | . | 3 prime UTR | RP1 | c.1625C>G | p.Ser542* | stop gained | 0.99940 |
| PHC1 | c.115-427G>T | . | intron | RP1 | c.1625C>G | p.Ser542* | stop gained | 0.99937 |
| BEST1 | c.*1721G>A; c.*1692G>A | .; . | 3 prime UTR; 3 prime UTR | TRPM1 | c.1879G>A | p.Val627Met | missense | 0.99937 |
| APC2 | c.1208-67_1208-66insGGGGGGGGG; c.967G>C | .; p.Gly323Arg | intron; missense | RP1 | c.1625C>G | p.Ser542* | stop gained | 0.99925 |
| TBR1 | c.1190+182_1190+184delTTT | . | intron | RP1 | c.1625C>G | p.Ser542* | stop gained | 0.99923 |
| TRPM1 | c.1879G>A | p.Val627Met | missense | RBP3 | c.2919C>A | p.Ser973Ser | synonymous | 0.99922 |
| RAD54B | c.945-1791_945-1789delAAA | . | intron | RP1 | c.1625C>G | p.Ser542* | stop gained | 0.99920 |
| ERCC1 | c.426-9C>T | . | intron | RP1 | c.1625C>G | p.Ser542* | stop gained | 0.99917 |

Table reports, for each digenic combination classified as pathogenic and for each gene included in the gene pair, the gene symbol and the HGVS of each variant referred to the specific gene (could be one heterozygous or homozygous variant, or two variants assumed in compound heterozygosity). A digenic combination is classified as pathogenic for the reported phenotypes if the associated score is greater than an optimized threshold of 0.13587435. If more than 50 digenic combinations are classified pathogenic by DIVAs, only the first 50 pathogenic ones are reported.

For Research Use Only. Not for use in diagnostic procedures.
